# A convolutional-recurrent neural network approach to resting-state EEG classification in Parkinson’s disease

**DOI:** 10.1101/2021.06.10.21258677

**Authors:** Soojin Lee, Ramy Hussein, Rabab Ward, Z. Jane Wang, Martin J. McKeown

## Abstract

**Background:** Parkinson’s disease (PD) is expected to become more common, particularly with an aging population. Diagnosis and monitoring of the disease typically rely on the laborious examination of physical symptoms by medical experts, which is necessarily limited and may not detect the prodromal stages of the disease.

**New Method:** We propose a lightweight (∼20K parameters) deep learning model, to discriminate between resting-state EEG recorded from people with PD and healthy controls. The proposed CRNN model consists of convolutional neural networks (CNN) and a recurrent neural network (RNN) with gated recurrent units (GRUs). The 1D CNN layers are designed to extract spatiotemporal features across EEG channels, which are subsequently supplied to the GRUs to discover temporal features pertinent to the classification.

**Results:** The CRNN model achieved 99.2% accuracy, 98.9% precision, and 99.4% recall in classifying PD from healthy controls (HC). Interrogating the model, we further demonstrate that the model is sensitive to dopaminergic medication effects and predominantly uses phase information of the EEG signals.

**Comparison with Existing Methods:** The CRNN model achieves superior performance compared to baseline machine learning methods and other recently proposed deep learning models.

**Conclusion:** The approach proposed in this study adequately extracts the spatial and temporal features in multi-channel EEG signals that enable the accurate differentiation between PD and HC. It has excellent potential for use as an oscillatory biomarker for assisting in the diagnosis and monitoring of people with PD. Future studies to further improve and validate the model’s performance in clinical practice are warranted.

## 1. Introduction

Parkinson’s disease (PD) is the second most common neurogenerative disorder next to Alzheimer’s disease, affecting 1–2% of people over age 65 years old (Scandalis et al., 2001). The estimated prevalence and incidence of people diagnosed with PD are expected to grow due to an increasingly aging population and greater recognition of the symptoms. According to a recent epidemiological study, individuals with PD more than doubled from 2.5 million in 1990 to 6.1 million in 2016 (Feigin et al., 2019). PD is a progressive disorder marked by the degeneration of dopaminergic neurons in the substantia nigra pars compacta projecting to the basal ganglia (BG). Although PD is most recognized for its cardinal motor symptoms of slowness of movement, tremor, rigidity, and postural instability, a broad spectrum of non-motor symptoms such as cognitive impairment, sleep disorders, and autonomic dysfunction are also frequently observed (Poewe et al., 2017).

Today, the diagnosis of PD is clinical, meaning that individuals with PD are diagnosed by a clinician if two or more cardinal symptoms are present. Unfortunately, no definitive single biomarker (e.g., a blood biomarker) that can accurately detect PD early. Accordingly, the diagnosis can be difficult when no significant physical signs or symptoms are yet present in individuals, such as those who have very early or prodromal PD. Early diagnosis is vital, since overt motor symptoms may only present when the majority of dopaminergic neurons are lost (Poewe et al., 2017). Any “neuroprotective” strategies would ideally be applied before then. Once diagnosed, disease progression is typically monitored using the United Parkinson’s Disease Rating Scale (UPDRS) by a medical expert in a clinical setting, which is generally done at intervals of 6–12 months. Limitations to this approach (Heijmans et al., 2019) include: 1) disease progression and symptoms are monitored infrequently, failing to comprehensively capture patients’ experiences that often vary over time between visits to a clinic, and 2) evaluating UPDRS is laborious, time-consuming, error-prone, and also partially subjective (Haaxma et al., 2008).

In the past years, efforts have been devoted to improving detection and monitoring of PD using data that are informative and relatively easy to collect from individuals with PD such as speech and handwriting (for review, see (Bind et al., 2015; Pereira et al., 2019; Thomas et al., 2017)). One investigative modality is electroencephalography (EEG), which has superb temporal resolution and relatively low cost. Compared to the well-characterized pathological oscillations of PD described in the subcortical structures (Brown et al., 2001; Priori et al., 2013), how PD influences neural oscillations in cortical regions remains relatively unclear. Several studies have reported PD is associated with slowing of cortical rhythms and resultant increased power of low-frequency rhythms (<10 Hz) in the occipital region (Geraedts et al., 2018; Morita et al., 2011; Neufeld et al., 1994; Soikkeli et al., 1991). However, slowing of the occipital oscillations is not specific to PD and has been commonly observed in people with other neurodegenerative conditions such as Alzheimer’s disease (Cassani et al., 2018).

Several recent studies have discovered that PD is related to a variety of linear and nonlinear changes in EEG signals, including abnormal patterns in the bispectrum (Yuvaraj et al., 2018), changes in entropy computed from wavelet packet decomposition coefficients over broad frequency ranges (Han et al., 2013), and increased coherence in frontal inter-hemispheric regions (Carmona et al., 2017), especially in the frequency range of 10–35 Hz (Silberstein et al., 2005). These studies indicate that the quantitative analysis of EEG can be potentially valuable for diagnosis, prognosis, monitoring, and planning of treatment strategies in PD. Quantitative EEG features extracted in the time domain, frequency domain, and time-frequency domain have all been utilized by a classification model based on machine learning to differentiate between PD and healthy individuals (Koch et al., 2019; Oliveira et al., 2020; Yuvaraj et al., 2018). These studies have provided valuable information by identifying the discriminatory EEG features that could be directly linked to the abnormal changes induced by the disease. However, the strategy of using hand-crafted features in classification may result in limited performance when tested on unseen data coming from new subjects (Cao et al., 2018).

Recent progress in deep learning has resulted in substantial advances in identifying, classifying, and quantifying distinguishable patterns in clinical data in various medical fields (Su et al., 2020; Woo et al., 2017). Deep learning models have been applied to PD EEG to perform a range of tasks such as discriminating on-medication from off-medication conditions (Shah et al., 2020) and classification between PD and healthy controls (HC) based on EEG collected during a specific task (Shi et al., 2019). However, there are only three studies (Lee et al., 2019a; Oh et al., 2018; Xu et al., 2020) that have deployed deep learning to discriminate between PD and HC using resting-state EEG. In (Oh et al., 2018), a thirteen-layer convolutional neural network (CNN) structure was proposed, which achieved 88.3% accuracy, 84.7% sensitivity, and 91.8% specificity using raw resting EEG data collected from 20 PD and 20 HC. The authors in (Xu et al., 2020) proposed a pooling-based recurrent neural network (RNN) consisting of several long short-term memory (LSTM) layers to extract both long- and short-term dependencies in EEG data that are crucial for EEG classification. The model was applied to the raw EEG data collected from 20 PD and 20 HC and achieved comparable performance as in (Oh et al., 2018) with 88.3% precision, 84.8% sensitivity, and 91.8% specificity. The third is our proof-of-concept work (Lee et al., 2019a) where we proposed a hybrid convolutional-recurrent neural network (CRNN) model based on CNN and LSTM. Although the model achieved a high accuracy of 96.9% in the PD and HC classification task, the study had a limitation in that comparison with other classification models was not done and it was not definitive that the result was achieved due to the model, the dataset or a combination of both.

In this work, we propose a hybrid CRNN model based on CNN and gated recurrent units (GRUs) to classify PD and HC individuals based on resting-state EEG, with the aim of achieving the following three objectives: 1) to develop a model that uses less memory and is computationally more efficient, 2) to achieve state-of-the-art classification performance, and 3) to conduct comprehensive evaluations of the proposed model by comparing its performance with the state-of-the-art CNN model in (Oh et al., 2018) and with those obtained by several traditional machine learning algorithms that have been widely used in prior studies. In addition, we investigated whether the CRNN model utilizes clinically relevant EEG features and conducted simulation studies to add interpretability to the model.

The remainder of this paper is organized as follows. Section 2 provides information on the study participants and EEG datasets, introduces the proposed model as well as other classification models to compare, and describes how the models were trained and evaluated. Experimental results are reported in Section 3, and we discuss the results and conclude this paper in Section 4.

## 2. Materials and methods

### 2.1. Participants

Twenty people with PD and 22 age- and sex-matched HC participated in this study. Participants with atypical parkinsonism or other neurological disorders were excluded from the study. All included PD participants were classified as having mild stage PD (Hoehn and Yahr Stage 1-2) and did not have drug-induced dyskinesia. One HC participant was excluded from data analysis due to severe muscle artifacts in the recorded EEG. Therefore, 20 PD (10 males; 10 females; age: 67.6 ± 7.0 years) and 21 HC (11 males; 10 females; age: 67.5 ± 6.4 years) participants were included in the analysis (Table 1). The study protocol was approved by the Clinical Research Ethics Board at the University of British Columbia (UBC), and the recruitment was conducted at the Pacific Parkinson’s Research Centre (PPRC) on the UBC campus. All participants gave written, informed consent before the experiment.

**Table 1.** Demographic and clinical characteristics of the study participants

|  | PD (n=20) | HC (n=21) |
| --- | --- | --- |
| Age (year) | $67.6 \pm 7.0$ | $67.5 \pm 6.4$ |
| Sex (male/female) | 10/10 | 11/10 |
| Disease duration (year) | $7.6 \pm 4.3$ | - |
| UPDRS III | $23.5 \pm 9.8$ | - |
| UPDRS III item 3.16 (kinetic tremor) | $2.3 \pm 1.3$ | - |
| UPDRS III item 3.17 (rest tremor) | $2.4 \pm 2.2$ | - |
| Hoehn and Yahr scale | 1–2 | - |
| Levodopa Equivalent Daily Dose (mg) | $708.2 \pm 389.9$ | - |

### 2.2. Study protocol

The participants were comfortably seated in front of a computer screen and were instructed to focus their gaze on a continuously displayed fixed target while EEG was being recorded. EEG was recorded for a fixed duration of 60 s. For the HC participants, the recording session was performed once, whereas there were two recording sessions for the PD participants: one in off-medication and the other in on-medication condition on the same day. The PD participants stopped taking their normal Levodopa (L-dopa) medication at least 12 hours, and any dopamine agonists 18 hours prior to the beginning of the experiment. UPDRS Part III (motor examination) was assessed in the off-medication condition. Immediately after finishing the first EEG acquisition, they took a regular dose of L-dopa medication and rested for one hour. After the break, EEG was recorded in the on-medication condition.

### 2.3. EEG recording and preprocessing

EEG was recorded from 27 scalp electrodes using a 64-channel EEG cap (Neuroscan, VA, United States) and a Neuroscan SynAmps2 acquisition system (Neuroscan, VA, United States) at a sampling rate of 1 kHz. Recording electrodes were positioned according to the International 10-20 placement standard, and two additional pairs of surface electrodes were used to detect vertical and horizontal eye movements. Impedances were kept below 15 kΩ using Electro-Gel (Electrode-Cap International, OH, United States). The EEG data were bandpass filtered between 1 and 55 Hz using a two-way finite impulse response (FIR) filter (the “eegfilt” function in EEGLAB) to remove DC offsets, slow drifts (< 1 Hz), and line noise at 60 Hz. Data were re-referenced to average reference, and stereotypical artifacts including ocular artifacts (EOG) and muscle tension were removed using an automatic artifact rejection method (Winkler et al., 2011) based on independent component analysis (ICA) (Makeig et al., 1996). The data were then standardized and segmented into 1-s or 2-s epochs (= samples) to train and validate the classification models described in the next sections. We segmented the data into smaller epochs to divide the non-stationary EEG signals into several pseudo-stationary segments expected to have comparable characteristics. In addition, segmentation has the advantage of generating a larger number of labeled EEG samples, which notably boosts the detection performance of the PD classification models.

### 2.4. Machine learning pipeline

We developed several models based on traditional machine learning algorithms to obtain baseline classification performance and compare them with the deep learning models introduced in section 2.5. Fig. 1A displays a block diagram showing the pipeline of developing traditional machine learning models. It consists of three main modules: feature extraction, feature ranking/selection, and classification. In comparison, Fig. 1B depicts a block diagram of the pipelines to develop deep learning-based models, absent of feature extraction and feature ranking/selection steps.

**Fig. 1.**
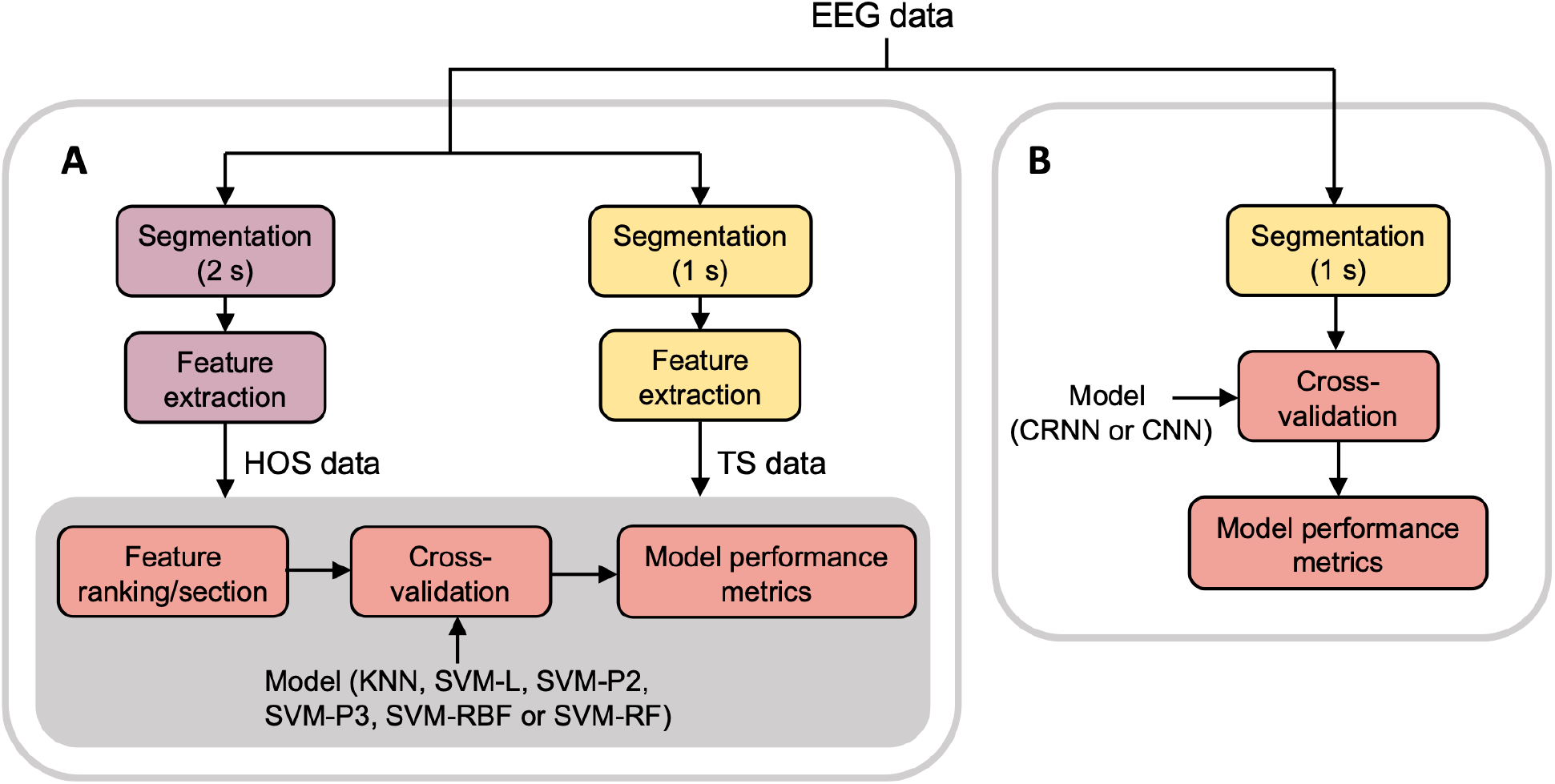
Block diagrams showing the overall processes of PD EEG classification based on **(A)** machine learning models and **(B)** deep learning models.

#### 2.4.1. Time-series feature extraction

The machine learning models were trained and validated on two datasets created from the same EEG data (Fig. 1). The first dataset (HOS dataset) was created based on the prior work (Yuvaraj et al., 2018) utilizing higher-order spectra (HOS) features to discriminate PD and HC resting-state EEG. The features were derived from the EEG bispectrum, a third-order analogue of the power spectrum that describes nonlinear interactions between oscillations (Mahmoodian et al., 2019). The continuous 60-s EEG signals were first segmented into 2-s epochs (Yuvaraj et al., 2018), and the bispectrum *B*(*f*_1_, *f*_2_) defined in Eq. (1) was computed for each EEG channel and epoch with 50% overlap Hanning window and 512 NFFT points (NFFT point is the number of discrete Fourier transform (DFT) points used to estimate power spectral density).

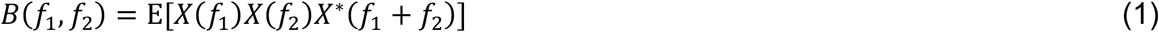

where *X*(*f*) is the DFT of the signal and *X*\*(*f*) is the conjugate of *X*(*f*).

Then, 13 features, including mean magnitude, bispectrum entropies, phase entropy, bispectrum moments, weighted center of bispectrum, and absolute weighted center of bispectrum were extracted (Yuvaraj et al., 2018). Accordingly, the HOS dataset contained 1230 samples in total (600 and 630 for PD and HC, respectively), each with 351 (= 13 × 27 EEG channels) bispectrum features extracted from the 27 channels.

The second dataset (TS dataset) was created based on the prior work by (Koch et al., 2019) where an automated feature extraction pipeline was used to extract 794 time-series features from the PD and HC resting-state EEG signals. We segmented the 60-s EEG signals into 1-s epochs and used the same *tsfresh* python package (Christ et al., 2018) with its default setting to extract 779 time-series features (e.g., autocorrelation, quantiles, number of zero-crossing) from each channel. This generated 21,033 (= 779 × 27 channels) features per epoch. In addition to the automatically generated time-series features, we considered the following 7 spectral features derived from the fast Fourier transform (Koch et al., 2019) that are associated with PD (Geraedts et al., 2018; Koch et al., 2019): peak frequency in the occipital channels (O1, Oz, O2) and mean theta (4–8 Hz), alpha (8–13 Hz), beta (13–30 Hz), and gamma (30–50 Hz) power across the 27 channels. As a result, the TS dataset included 2,460 samples (1,200 and 1,260 for PD and HC, respectively), each with 21,040 features.

The two datasets were then used to train and test several machine learning models after feature ranking and selection processes.

#### 2.4.2. Feature ranking and selection

Feature selection plays a crucial role in machine learning in that it removes redundancy, reduces computational time, and improves the predictive performance of the classification algorithms (Awan et al., 2019). Therefore, we performed feature ranking and selection to identify a subset of the features with the most significant impact on PD recognition accuracy. We used univariate statistical analysis (ANOVA) and identified significant features with a significance level of *α* = 0.05. Then, either all or a subset of the significant features were used to train the machine learning models. Selecting a subset of the features was done by ranking the retained features based on the ANOVA p-values, selecting the top 25 features, and then gradually adding the next 25 significant features.

#### 2.4.3. Machine learning models

We considered the following machine learning algorithms that have been most commonly implemented in prior EEG studies on PD and HC classification (Maitín et al., 2020):

- *k*-nearest neighbor (KNN)
- Support vector machine (SVM) with a linear kernel (SVM-L)
- SVM with a polynomial kernel with degree of 2 (SVM-P2)
- SVM with a polynomial kernel with degree of 3 (SVM-P3)
- SVM with a radial basis function (RBF) kernel (SVM-RBF)
- Random forest (RF)

The KNN algorithm assigns a class to the current feature vector based on its *k* nearest neighbors. SVM is a supervised machine learning algorithm based on statistical learning theory to solve binary/multiclass classification or regression problems. For a binary classification problem, linear SVM makes a simple linear separation between the two classes by finding the maximum-margin hyperplane. In contrast, the polynomial kernel makes the linear decision boundary more complex as the degree of the kernels increases and can handle non-linearly separable data (Noble, 2006). The RBF kernel, also known as Gaussian kernels, finds the nonlinear decision boundary by mapping input data into a high dimensional feature space through the RBF kernel function. A RF classifier is an ensemble of several decision tree classifiers (Breiman, 2001), whereby bootstrapping is applied to a subset of the available features, a decision tree classifier is trained on the feature subset, and many different decision trees are obtained by repeating the process. The final decision is made by aggregating the decisions of individual trees and taking the majority vote.

The HOS and TS datasets were used to train the above six classifiers. Each classifier contains hyperparameters that need to be tuned to achieve reasonable classification results. The hyperparameters and search space under consideration are listed in Table 2. There are various methods to optimize the hyperparameter values, and we chose the grid search and random search methods (Bergstra and Bengio, 2012) (Table 2). The hyperparameter optimization procedure was applied with a 5-fold cross-validation (CV) procedure, whereby the models were trained with different hyperparameters on 4-folds of the training set and the best hyperparameters were determined based on the model accuracy on the remaining 1-fold of the training set. Note that we adopted a nested CV approach (see section 2.6) and the hyperparameter optimization was done in the inner loop CV (Fig. 2).

**Table 2.** Hyperparameter search space and search method for the six classifiers

| Classifier | Search method | Hyperparameter | Space |
| --- | --- | --- | --- |
| KNN | Grid search | Number of neighbors | [6, 8, 10, 12, 14, 16, 18, 20] |
| SVM-L | Grid search | Soft margin constant | [1, 10, 100] |
| SPM-P1 | Grid search | Soft margin constant | [1, 10, 100] |
| SPM-P2 | Grid search | Soft margin constant | [1, 10, 100] |
| SPM-RBF | Grid search | Soft margin constant<br>RBF kernel parameter gamma | [1, 10, 100]<br>[0.001, 0.01, 0.1, 1] |
| RF | Random search | Number of trees<br>Max depth of each tree<br>Min number of samples required to split a node<br>Min number of samples required in the leaf node | [5, 6, 7, ..., 200]<br>[5, 6, 7, ..., 50]<br>[2, 3, 4, ..., 20]<br>[1, 2, 3, ..., 10] |

**Fig. 2.**
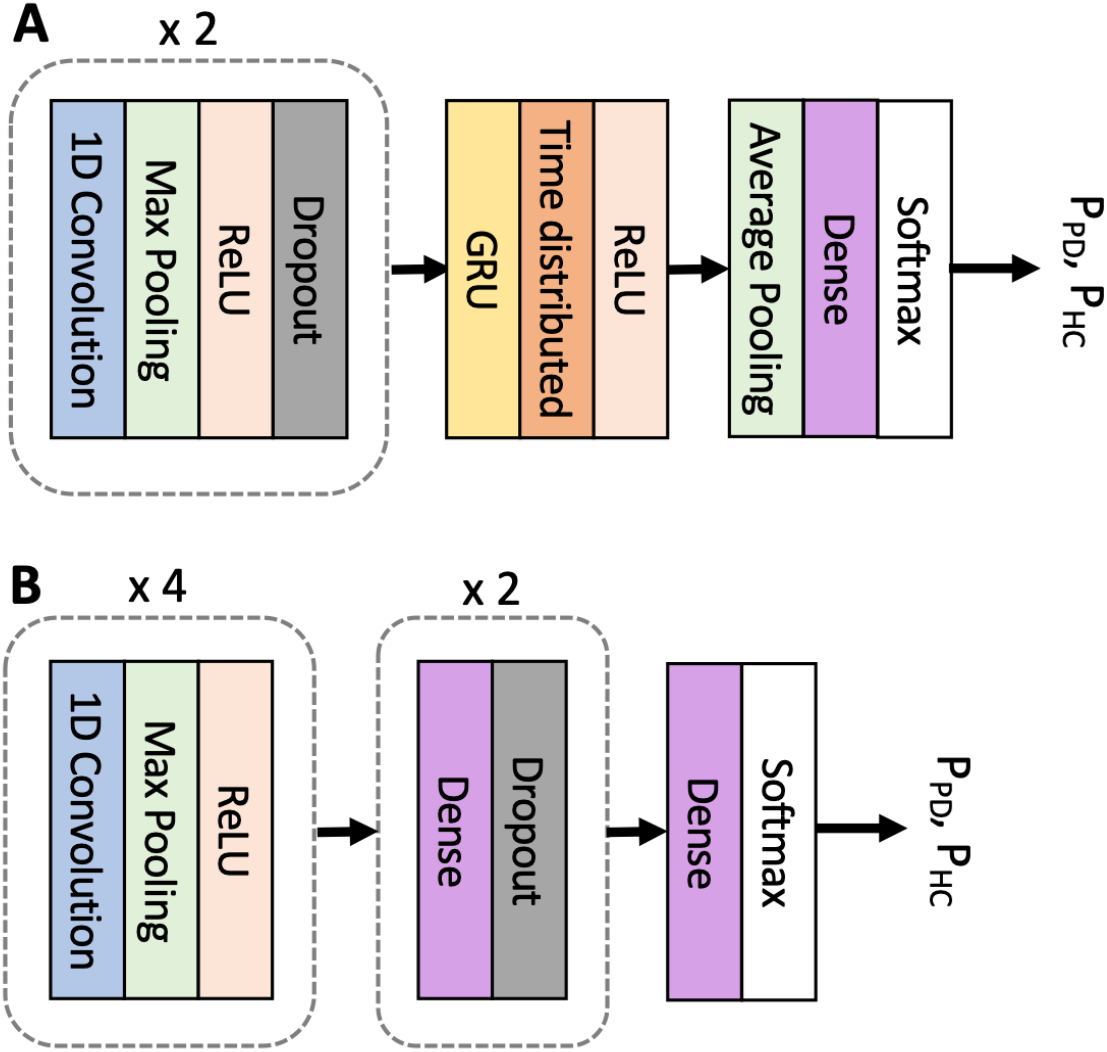
Nested CV used to evaluate classification models under study. The dataset is first split into train and test sets in the outer loop. The outer train set in each fold is used to select the model hyperparameters that maximize the inner CV accuracy. The best hyperparameters are then used to train the model using the entire samples in the outer train set, which is then tested on the outer test set to obtain 1 out of K (=10) performance scores (accuracy, precision, recall, F1 score, and AUC). The average of the performance scores over K folds is reported as the final results.

### 2.5. Deep learning pipeline

#### 2.5.1. Data preparation

We used non-overlapping 1-s epochs as input data for the deep learning models. The epoch length is the same as the one used to create the TS dataset (see Section 2.4.1 and Fig. 1A), and therefore it enables a fair comparison of the classification performance between the machine learning and deep learning models without introducing any potential biases that arise from different epoch lengths and number of samples. The raw EEG signals were directly used as input data without additional feature extraction or feature selection processes.

#### 2.5.2. Deep learning models

In this section, we provide an overview of the proposed CRNN model (Fig. 3A) and a benchmark CNN model proposed in (Oh et al., 2018) (Fig. 3B). The CNN model was adopted in this study for a comparison with our model.

**Fig. 3.**
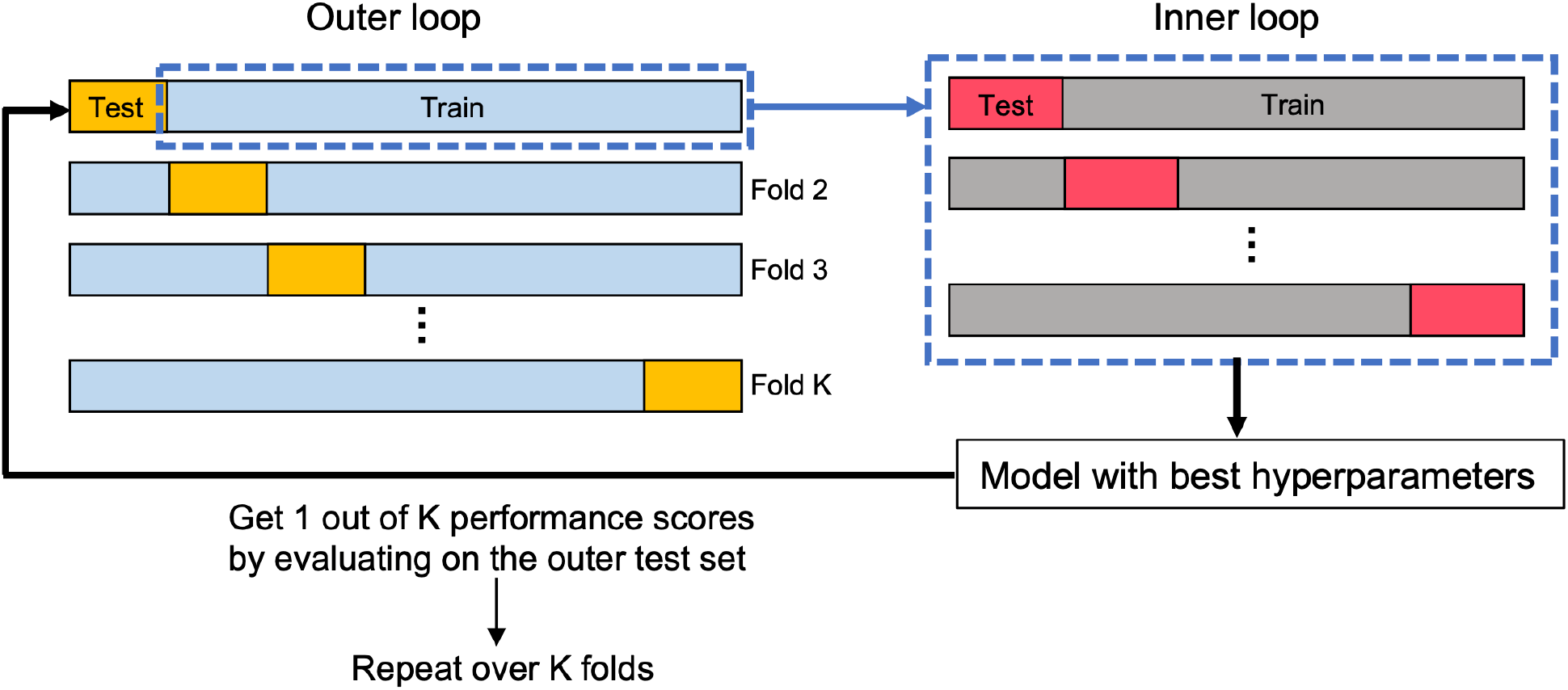
Illustration of the core architecture of the neural network models. Both models take 1-s 2D EEG timeseries (time points × channels) as input and generate the probability of each class (*P*_*PD*_ and *P*_*HC*_) as ouput. **(A)** Proposed CRNN model. It contains 2 consecutive blocks consisting of a 1D convolutional layer, a Max Polling layer, a ReLU activation, and a Dropout layer. The 3^rd^ block consists of a GRU layer, a time distributed layer, and a ReLU activation. The 4^th^ block contains an average pooling layer, a dense layer, and a softmax layer. **(B)** CNN model. It contains four consecutive blocks consisting of a 1D convolutional layer, a Max Pooling layer, and a ReLU activation. The 5^th^ and 6^th^ blocks consist of a dense layer and a Dropout layer. The 7^th^ block consists of a dense layer and a softmax layer.

The CRNN model relies on 1D CNN and GRU cells to effectively capture both spatial and temporal features in multi-channel EEG signals. The dimensions of the input data to the CRNN model are *d* × *N*_*ch*_ (= 1000 × 27), where *d* is the number of time points in each sample and *N*_*ch*_ is the number of EEG channels. The total number of EEG samples (*N*) is 2460 (= 60 samples/participant × 41 participants). Two 1D-convolutional layers are first used to extract spatiotemporal features from the EEG signals. In these layers, 1D kernels of height *k* (covering *k* time points) and width 27 (covering the 27 channels) slide across the EEG samples in the time domain. The output of the convolutional layers is then fed into an RNN with GRU cells to extract temporal dependencies dwelling in the extracted features. Finally, two fully connected layers (time distributed and dense) are used on top of the RNN layer to transform the extracted high-level features into more abstract features that are used for reliable EEG classification. A max-pooling layer is used after each 1D-convolutional layer to downsample the feature maps to reduce the number of trainable parameters and computation cost. Dropout layers are also adopted to prevent the model from overfitting. We used a ReLU activation function for all layers and a softmax activation function for the final layer. The CRNN model was trained using the Adam optimizer (Kingma and Ba, 2015) with a learning rate of 0.001 to optimize the softmax binary cross-entropy loss. The hyperparameter search spaces were [3, 7, 11, 15], [16 32, 64], [20, 25, 30, 35] and [25, 35, 45] for the kernel sizes, number of convolutional filters, number of GRU units, and number of dense units, respectively.

For a comparison with the CRNN model, we adopted the CNN model recently proposed in (Oh et al., 2018). This model includes 4 consecutive blocks, each consisting of a 1D-convolutional layer, a max-pooling layer, and a ReLU activation layer, and three fully connected layers are built on top of the four CNN-based blocks (Fig. 3B). The idea behind the architecture is that the first convolutional layer generates feature maps from the input data using 1D-convolution kernels, and the feature maps are subsequently used as input data for the upper convolutional layers, resulting in the deeper layers to learn higher representations that are relevant to the classification problem. Similar to the CRNN model, the CNN model was trained with Adam optimizer with a learning rate of 0.001 to optimize the binary cross-entropy cost function. A ReLU activation function was used in all layers except for the final layer where a softmax activation was used. The hyperparameter search spaces were [5, 9, 13, 17], [4, 8, 12, 16, 20], [25, 30, 35], and [5, 10, 15] for the kernel sizes, number of filters, number of units in the first dense layer, and number of units in the second dense layer, respectively.

The batch size and number of epochs to train CRNN and CNN models were the same as 64 and 100, respectively. All the hyperparameters were tuned using the grid search optimization technique with the search spaces described above, and the final values were set to those that achieved the best training accuracy. The optimal set of hyperparameters for the CRNN model was found to be 3, 35, and 35 for the kernel size, number of GRU units, and number of units in the dense layer, and the number of filters in the convolutional layers were 32 and 64 (Table 3). For the CNN model, the optimal set of hyperparameters were found to be 9 for the kernel size, 8, 12, 12, and 16 for the number of filters in the convolutional layers, and 30 and 5 for the number of units in the dense layers (Table 4). The final architecture and parameters of the two models are shown in Tables 3 and 4.

**Table 3.** Architecture and parameters of the CRNN model

| Layer | Number of filters | Kernel size | Stride | Output shape | Number of parameters | Regularization |
| --- | --- | --- | --- | --- | --- | --- |
| Input | - | - | - | (1000, 27) | 0 | - |
| 1D convolution | 32 | 3 | 1 | (998, 32) | 2624 | - |
| Max-pooling | 32 | 3 | 2 | (499, 32) | 0 | Dropout (0.5) |
| 1D convolution | 64 | 3 | 1 | (497, 64) | 6208 | - |
| Max-pooling | 64 | 3 | 2 | (248, 64) | 0 | Dropout (0.5) |
| GRU | 35 | - | - | (248, 35) | 10500 | - |
| Time distributed | 35 | - | - | (248, 35) | 1260 | - |
| Global average pooling | 35 | - | - | (1, 35) | 0 | - |
| Dense | 35 | - | - | (1, 2) | 72 | - |
| Total |  |  |  |  | 20664 |  |

**Table 4.** Architecture and parameters of the CNN model

| Layer | Number of filters | Kernel size | Stride | Output shape | Number of parameters | Regularization |
| --- | --- | --- | --- | --- | --- | --- |
| Input | - | - | - | (1000, 27) | 0 | - |
| 1D convolution | 8 | 9 | 1 | (992, 8) | 1952 | - |
| Max-pooling | 8 | 2 | 2 | (496, 8) | 0 | - |
| 1D convolution | 12 | 9 | 1 | (488, 12) | 876 | - |
| Max-pooling | 12 | 2 | 2 | (244, 12) | 0 | - |
| 1D convolution | 12 | 9 | 1 | (236, 12) | 1380 | - |
| Max-pooling | 12 | 2 | 2 | (118, 12) | 0 | - |
| 1D convolution | 16 | 9 | 1 | (110, 16) | 1744 | - |
| Max-pooling | 16 | 2 | 2 | (55, 16) | 0 | - |
| Dense | - | - | - | (1, 30) | 26430 | Dropout (0.5) |
| Dense | - | - | - | (1, 5) | 155 | Dropout (0.5) |
| Dense | - | - | - | (1, 2) | 12 | - |
| Total |  |  |  |  | 32477 |  |

Our implementation was derived in Python using Keras with TensorFlow backend and underwent approximately 14 minutes of training on an NVIDIA TU102 GPU machine for the CRNN or CNN model. Once trained, the model took less than a second to test a new unseen data sample.

### 2.6. Performance evaluation

Accuracy, precision, recall, F1 score, and the area under the ROC curve (AUC) were used to assess classification performance of the classifiers. They were defined as

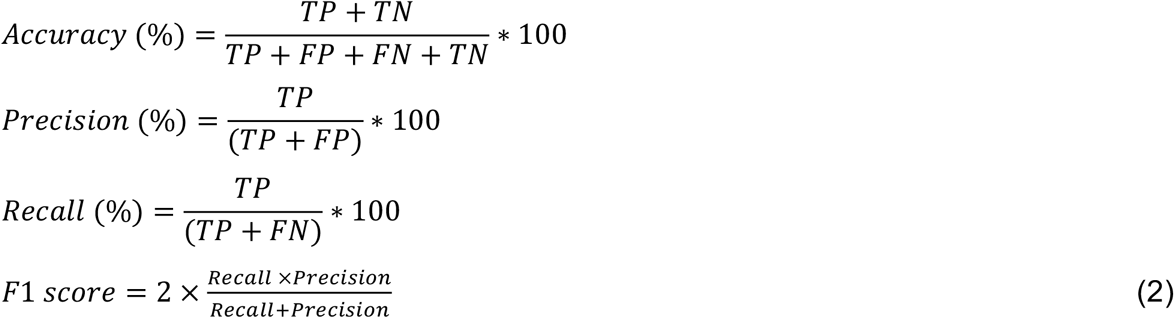

where *TP, TN, FP* and *FN* denote true positive, true negative, false positive, and false negative, respectively. *TP* is defined as the case where a PD sample is correctly classified. To overcome the bias in evaluating model performance, we adopted a nested CV strategy to train a model with hyperparameter optimization in the inner 5-fold CV and obtain an unbiased estimate of the expected accuracy of the model with the outer 10-fold CV (Wainer and Cawley, 2018) (Fig. 2). In the nested CV, the dataset is first split into a training set (90% of the total samples) to train the model and a test set (10% of the total samples) to provide the model accuracy estimate. In each fold of the outer CV, the training set is further used to tune the hyperparameters of the model to maximize the inner CV accuracy. The model with the best hyperparameters determined in the inner CV is trained using the entire samples in the outer training set and then tested on the outer test set to compute the performance metrics. This procedure is repeated 10 times through the outer 10-fold CV. We averaged the performance metrics obtained in each outer fold and reported them as the final results.

## 3. Results and discussion

### 3.1. PD classification performance

For the HOS dataset, the accuracy of the KNN, SVM-L, SVM-P2, SVM-P3, SVM-RBF, and RF models were in the range of 71.7–89.7%, 66.7–82.3%, 64.2–86.7%, 62.2–87.8%, 76.0–92.9%, and 71.4–85.4%, respectively, depending on the number of the features selected in the feature ranking and selection step (Supplementary Fig. S1A). Taking the SVM-RBF model as an example, it reached the maximum of 92.9% accuracy when the top 150 features were used, but adding more features resulted in inferior performance. Table 5 shows the best performance of the models when trained and tested on the HOS dataset along with the number of the selected features. The SVM-RBF performed the best, achieving 92.9% accuracy, 92.4% precision, 93.0% recall, 0.926 F1-score, and 0.928 ROC score. The KNN model was the second-best model, achieving 89.7% accuracy, 90.0% precision, 88.5% recall, 0.892 F1-score, and 0.897 ROC score.

**Table 5.** Performance of different classifiers (average 10-fold CV)

|  | Model | Accuracy (%) | Precision (%) | Recall (%) | F1 score | AUC | # of features |
| --- | --- | --- | --- | --- | --- | --- | --- |
| HOS data | KNN | 89.7 | 90.0 | 88.5 | 0.892 | 0.897 | 125 |
|  | SVM-L | 82.3 | 81.7 | 81.8 | 0.817 | 0.822 | 200 |
|  | SVM-P2 | 86.7 | 88.2 | 84.1 | 0.861 | 0.866 | 200 |
|  | SVM-P3 | 87.8 | 88.2 | 86.5 | 0.872 | 0.878 | 225 |
|  | SVM-RBF | **92.9** | **92.4** | **93.0** | **0.926** | **0.928** | 150 |
|  | RF | 85.4 | 87.5 | 81.9 | 0.843 | 0.854 | 175 |
| TS data | KNN | 92.6 | 90.4 | 94.8 | 0.925 | 0.927 | 2518 |
|  | SVM-L | 81.0 | 80.4 | 81.1 | 0.806 | 0.810 | 975 |
|  | SVM-P2 | 93.1 | 92.1 | 94.0 | 0.930 | 0.931 | 875 |
|  | SVM-P3 | 93.2 | 92.0 | 94.5 | 0.932 | 0.932 | 950 |
|  | SVM-RBF | **95.4** | **96.3** | **94.3** | **0.953** | **0.954** | 950 |
|  | RF | 87.8 | 89.0 | 85.5 | 0.872 | 0.877 | 800 |
|  | Proposed | **99.2** | **98.9** | **99.4** | **0.992** | **0.992** | - |
|  | CNN (Oh et al., 2018) | 95.4 | 95.2 | 95.5 | 0.953 | 0.954 | - |

When the TS dataset was used, the accuracy of the KNN, SVM-L, SVM-P2, SVM-P3, SVM-RBF, and RF models were in the range of 69.4–92.6%, 66.5–81.0%, 62.9–93.1%, 69.0–93.2%, 71.6–95.4%, and 70.7–87.8%, respectively, depending on the number of selected features. Supplementary Fig. S1B shows that adding more features up to 125 features resulted in a steep increase in the accuracy for all the models under study. The accuracy gradually improved up to 1000 features for SVM-RBF, SVM-P3, SVM-P2, and KNN whereas it remained relatively stagnant for the RF model. The best performance of each model and number of selected features are shown in Table 5. Compared to the HOS dataset results, all the models achieved better classification performance, suggesting that the features of the TS dataset are more representative than those of the HOS dataset. SVM-RBF was found to achieve the best classification performance with 95.4% accuracy, 96.3% precision, 94.3% recall, 0.953 F1-score, and 0.954 ROC score, followed by SVM-P3 and SVMP2 achieving similar accuracy scores of 93.2% and 93.1%, respectively.

Table 5 shows the classification performance of the CRNN and CNN models. The CNN model achieved 95.4% accuracy, 95.2% precision, 95.5% recall, 0.953 F1-score, and 0.954 ROC score, which were comparable to the performance of the SVM-RBF model when the TS dataset was used. The proposed CRNN model achieved 99.2% accuracy, 98.9% precision, 99.4% recall, 0.992 F1-score, and 0.992 ROC score, outperforming the CNN model with significant margins. This demonstrates the advantages of introducing a hybrid convolutional-recurrent neural network model in learning spatiotemporal EEG features needed for accurate EEG classification. It also shows the capability of the GRU in capturing abnormal temporal dynamics in EEG signals related to PD. In addition, contrary to the machine learning models that rely on hand-engineered features, our model has the advantage that feature learning is done automatically, relaxing the need for additional data processing steps (feature extraction, ranking, and selection).

### 3.2. Clinical relevance of the CRNN model classification

In the previous section, we demonstrated that the proposed CRNN model achieved state-of-the-art PD prediction using resting-state EEG data. After finalizing the model by training it on all the PD and HC samples, we probed the model to determine whether it could detect EEG features related to some clinical features of the PD participants. Specifically, we investigated whether 1) the probability of the EEG samples to be classified as PD (*P*_*PD*_) was related to the UPDRS part III score, and 2) the model classified more EEG samples as HC when the PD participants are on-medication status.

We first used the final CRNN model to estimate the PD probability for every EEG sample of the PD and HC participants and then computed individual-wise *P*_*PD*_ by averaging the probability across the 60 samples for each participant 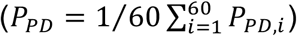. Fig. 4A shows a clear difference in the individual-wise *P*_*PD*_ between the PD and HC participants where the probability was close to one for the PD participants (1.00 ± 0.0013) and close to zero for the HC participants (0.00 ± 0.06). There was a significant correlation between the *P*_*PD*_ and UPDRS part III score (Fig. 4B: R = 0.53, p = 0.017), but the variation of the individual-wise *P*_*PD*_ across the UPDRS scores (*P*_*PD*_ range: 0.995–1) was very small. Note that the UPDRS score was not measured for HC participants.

**Fig. 2.**
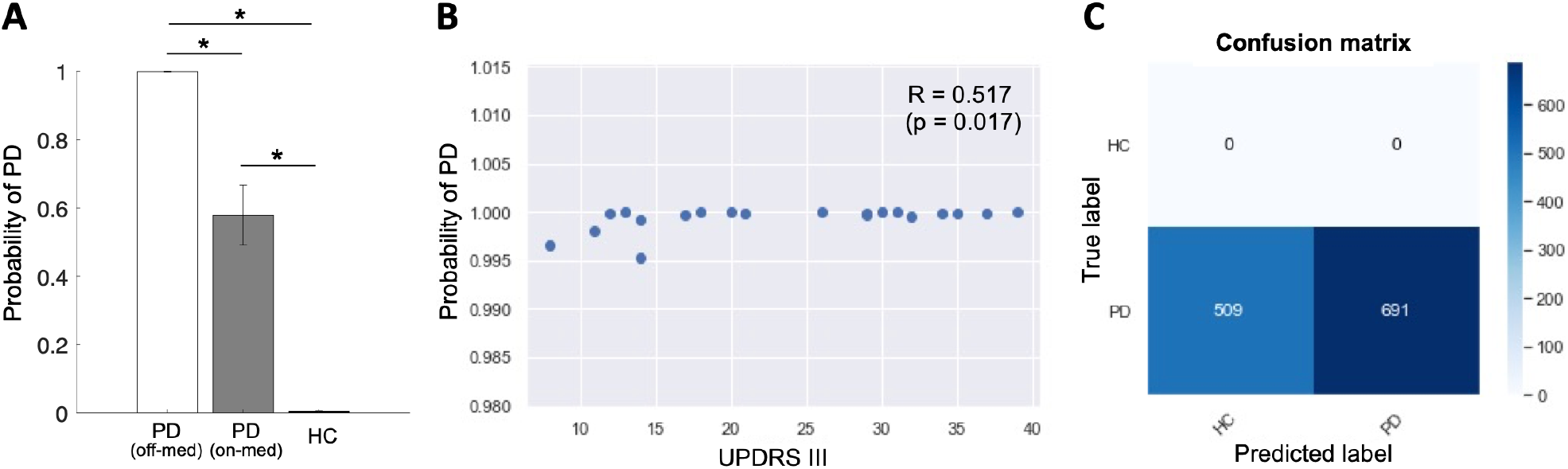
**(A)** The probability of PD (*P*_*PD*_) estimated by the CRNN model for the PD (off- and on-medication) and HC groups. The bar graphs shows the mean and standard deviation of *P*_*PD*_ across the participants in each group. P-values from the one-sample (off-vs. on-med) t-test and two-sample t-tests (PD off-med vs. HC; PD on-med vs. HC) are indicated with asterisks (*: Bonferroni-corrected p-value < 0.001). **(B)** A scatter plot of the *P*_*PD*_ and UPDRS part III scores of the PD participants demonstrates a positive correlation (R = 0.527, P = 0.017). **(C)** Confusion matrix showing that the CRNN model predicts 509 out of 1200 PD EEG samples as HC when the EEG was recorded an hour after the participants took a regular dose of L-dopa medication.

To investigate the influence of L-dopa medication on the classification performance, we used the EEG data recorded from the same PD participants when they were on medication. As the EEG data were recorded an hour after the participants took their regular L-dopa dose, the medication effects were expected to stay optimal at the time of recording. The middle bar in Fig. 4A indicates the individual-wise *P*_*PD*_ for the medicated PD participants (0.58 ± 0.39). The individual-wise probabilities significantly decreased compared to when the participants were off medication (one-sample t-test: *t*(19) = 4.80, p < 0.001) while the probabilities remained significantly higher compared to the HC group (two-sample t-test: *t*(39) = 6.72, p < 0.001). The decrease by medication was observed in 19 out of the 20 PD participants, indicating that for most of the participants the classification accuracy was reduced by the medication. Fig. 4C shows the confusion matrix of the predicted labels of the EEG samples (on-med PD). 509 out of the 1200 (42.4%) EEG samples were classified as HC, indicating that the model was able to detect that the EEG signals had become less pathological after the medication.

### 3.3. CRNN model inference in a simulation study

The CRNN model has the advantage that it achieves high classification accuracy by utilizing complex nonlinear temporal and spatial features in the EEG data. However, it is difficult to fully understand the nature of the CRNN model due to a lack of techniques available to explain and interpret results derived using deep learning algorithms. To gain a better understanding of what EEG features are learnt by the model, we conducted several simulation studies where we created simulated EEG datasets by manipulating spectral features of the original EEG data and examined the resultant changes in the prediction outcomes. Specifically, we manipulated the phases and power spectra of the EEG data, which are the most intensively investigated features in EEG studies.

#### 3.3.1. Effects of the randomized phases of EEG

In the first simulation study, we investigated the importance of EEG phases in model classification performance. We created a simulated EEG dataset by randomizing the phases but preserving the spectral powers of the original EEG data of the PD and HC participants. The model predicted 100% of the simulated EEG samples as HC regardless of their true labels, suggesting that it recognizes more randomized EEG phases as features related to HC. Interestingly, when we temporally reversed the EEG signals instead of randomizing the phases, the model predicted PD and HC samples correctly (accuracy = 99.2%). This suggests that the classification results are not merely based on the order of time points, but the model utilizes the phase information of the different frequency components of the EEG signals.

To investigate whether the effects of randomized phases were driven by a particular brain region, we randomized the phase of one EEG channel at a time and observed how this affected the *P*_*PD*_ estimated by the model. Then, we repeated the process across the 27 EEG channels. Fig. 5 shows the distribution of the estimated *P*_*PD*_ of the entire samples for the PD (N = 1,200) and HC (N = 1,260) groups. For the PD group, the *P*_*PD*_ was close to 1 only when the phases of the EEG signals in the left temporal and right frontal regions (channels 3, 4, 8, 9, 10, 11, 16, and 24) were randomized. The randomized phases of the other channels resulted the *P*_*PD*_ close to zero (i.e., HC). Similar results were obtained from the HC group that the randomized phases resulted the *P*_*PD*_ close to 0 except for the left temporal and right frontal regions. Taken together, the results suggest that the model learns more randomized EEG phases in the left temporal and right frontal regions and less randomized EEG phases in the other regions as PD-related features.

**Fig. 3.**
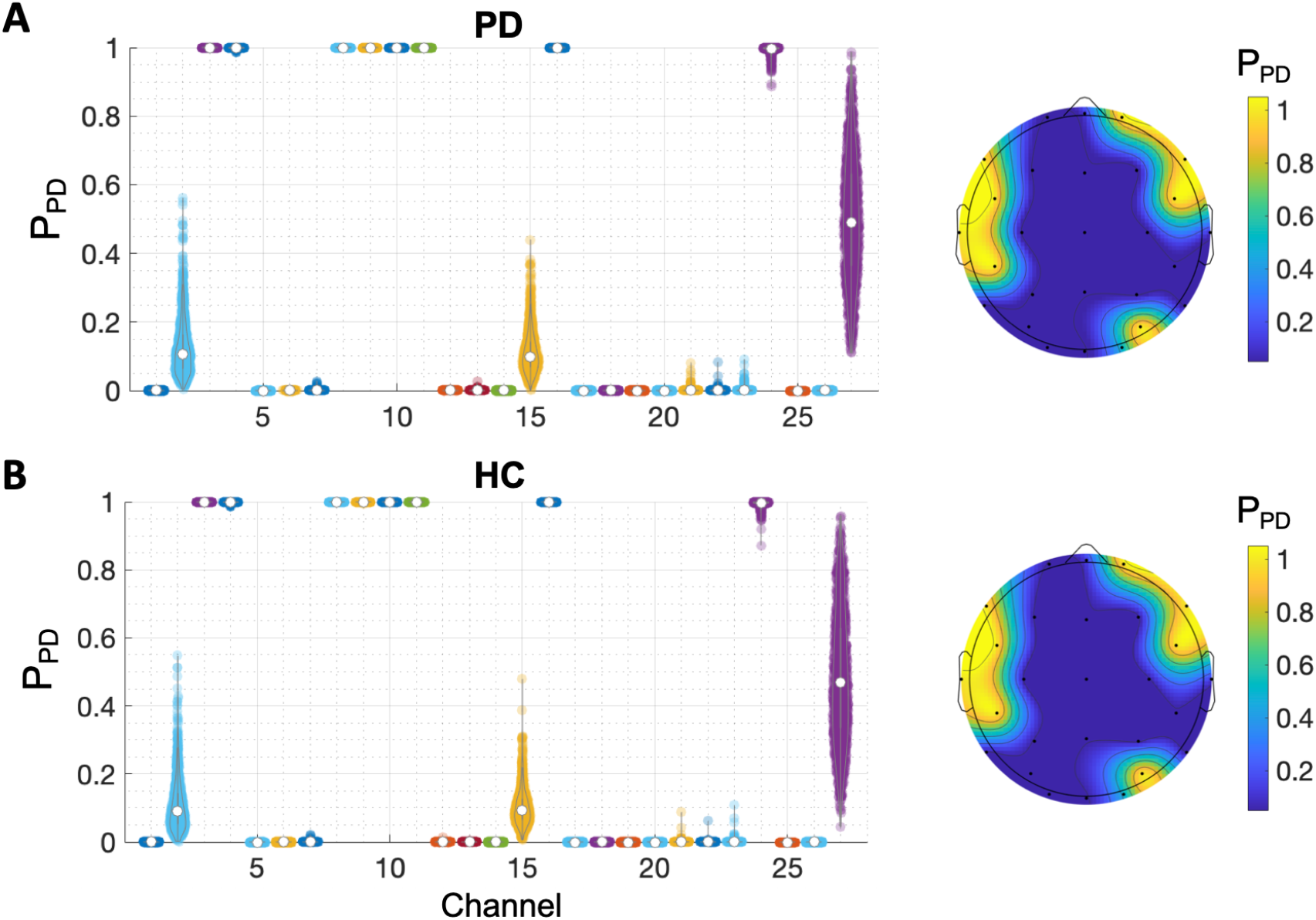
The distribution of *P*_*PD*_ of the EEG samples when the phase of one EEG channel was randomized. Higher *P*_*PD*_ indicates the sample is more likely from PD. The average values of the distributions are presented in the topographical scalp maps on the right. **(A)** Results from the PD EEG samples (N = 1200). **(B)** Results from the HC EEG samples (N = 1260).

#### 3.3.2. Effects of the spectral power of EEG

One of the most widely used methods to analyze EEG signals is power spectral analysis, which investigates spectral power of EEG signals in five historically pre-defined frequency bands (delta: 1-4 Hz; theta: 4-8 Hz; alpha: 8-13 Hz; beta: 13-30 Hz; gamma: > 30 Hz) that are linked to different functional characteristics of the brain (Groppe et al., 2013; Lee et al., 2019b). In the second simulation study, we investigated whether changes in the spectral power in each frequency band affected the model’s classification performance. For every electrode, we varied the spectral power of one of the frequency bands from - 100% (no power) to 100% (2 × the original power) while preserving the phase information intact. Fig. 6 shows the distributions of the *P*_*PD*_ of the EEG samples estimated by the model when the spectral power changes occurred. The *P*_*PD*_ was consistently estimated close to 1 for most of the EEG samples of the PD participants (Fig. 6A). Similarly, the model accurately predicted the *P*_*PD*_ of the HC samples close to 0 despite the spectral power changes (Fig. 6B).

**Fig. 6.**
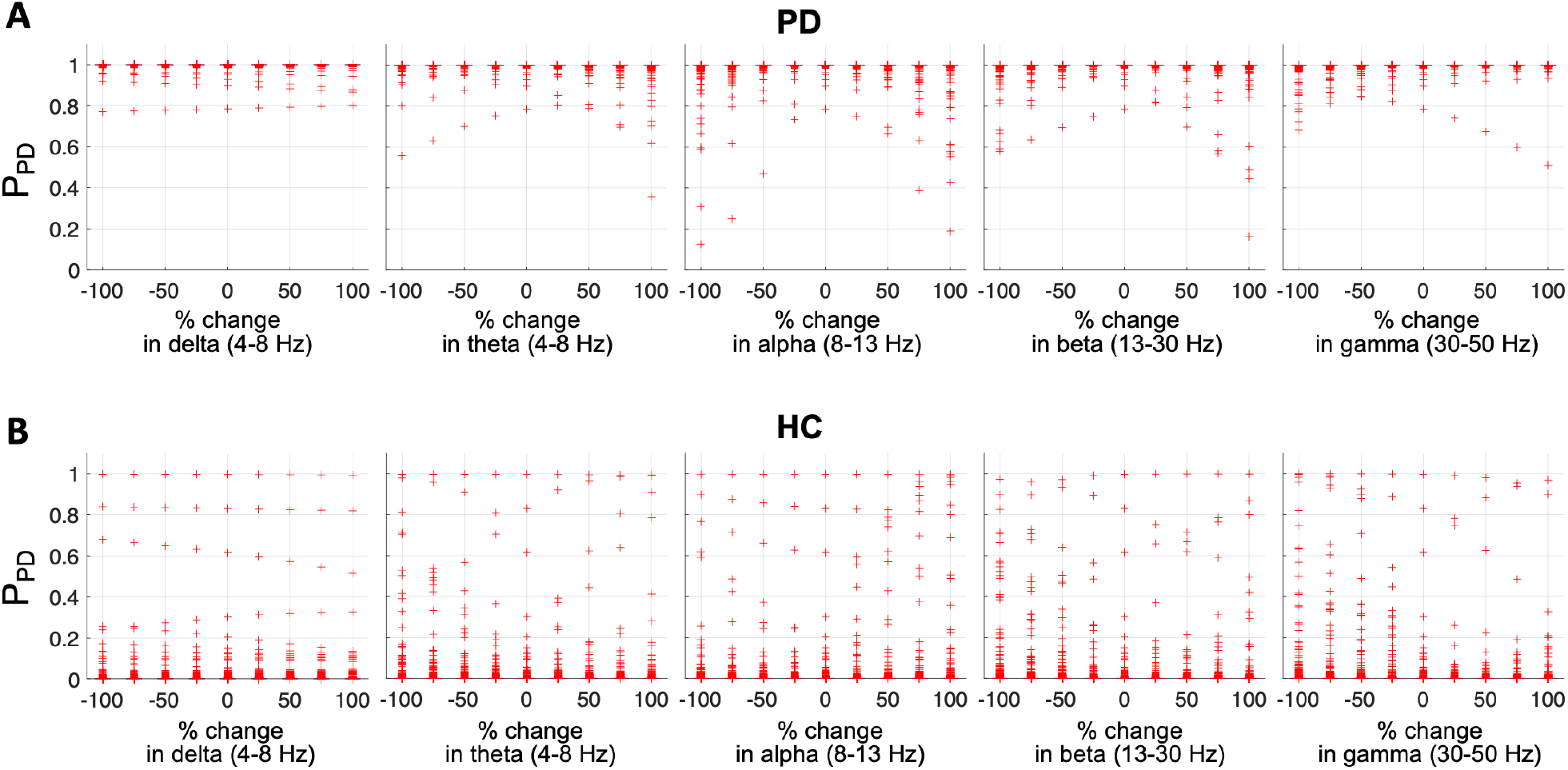
Effects of global spectral power change on the CRNN model performance. The spectral power in the delta, theta, alpha, beta or gamma frequency band was changed at every electrode by -100% (equivalent to zero power) to 100% (equivalent to doubled power). **(A)** *P*_*PD*_ of the PD EEG samples estimated by the model. **(B)** *P*_*PD*_ of the HC EEG samples estimated by the model.

We investigated whether a regional spectral power change instead of the global changes has effects on the model performance. To this end, the spectral band power was varied at each electrode at a time while keeping phase information the same. Fig. 7 shows topoplots where the value at each electrode represents mean *P*_*PD*_ obtained by averaging the estimated *P*_*PD*_ of the entire PD (first row) and HC (second row) samples when the spectral power at the electrode increased by 100%. For example, *P*_*PD*_ = 0.998 at F3 denoted in the figure indicates that the model estimated *P*_*PD*_ of the PD EEG samples to be 0.998 on average when the delta power at F3 increased by 100%. The topoplots show that the regional changes in the spectral power have little effect on the model classification performance for both PD and HC EEG data.

**Fig. 7.**
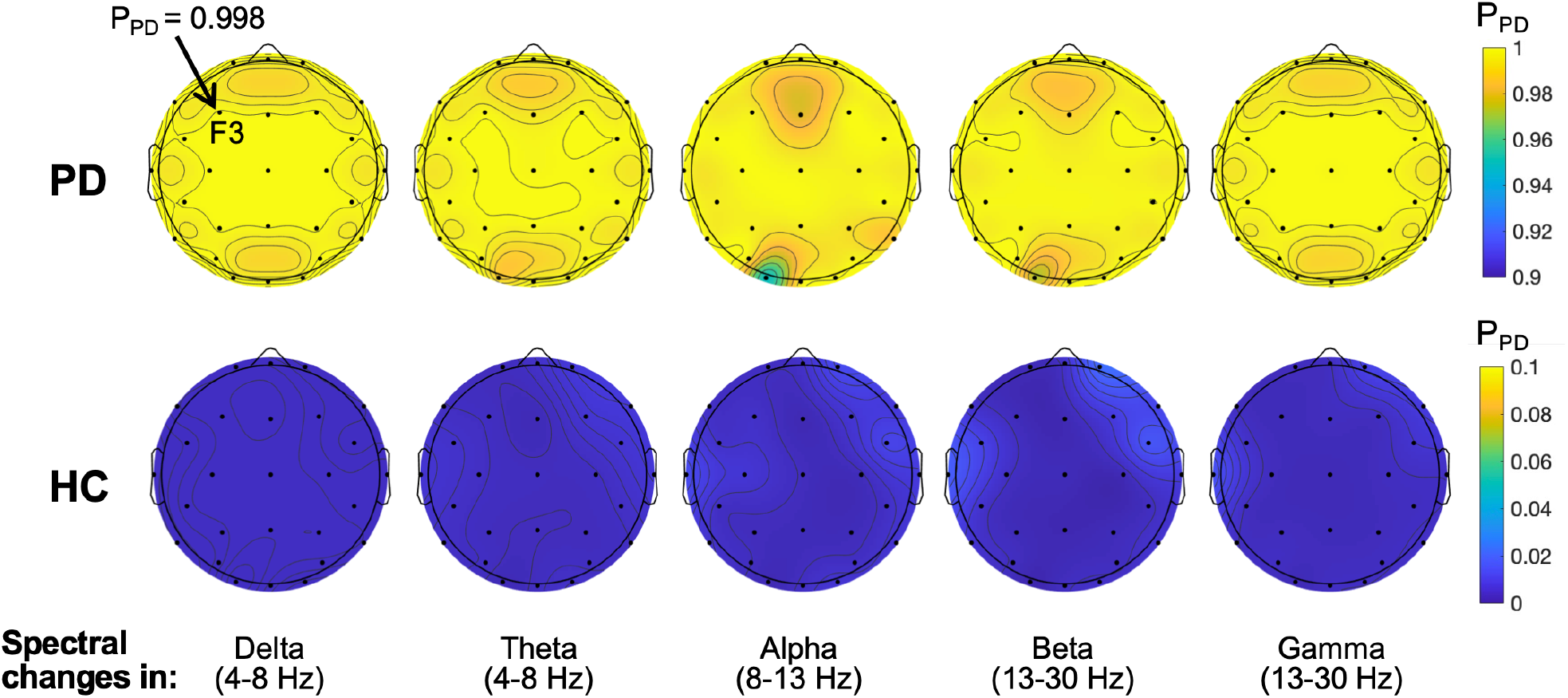
Effects of regional spectral power change on the CRNN model performance. The spectral power in the delta, theta, alpha, beta or gamma frequency band was changed at each electrode at a time by 100%, and *P*_*PD*_ of the EEG samples were estimated by the model. The mean *P*_*PD*_ across the EEG samples is presented in each topoplot with the colour bar on the righthand side showing the probability scale. The topoplots in the first and second rows demonstrate the results from the PD and HC EEG samples, respectively.

These results suggest that spectral characteristics of the EEG data are not the primary features used in the CRNN model.

#### 3.3.3. Effects of phase and spectral power of EEG

In practice, it is unlikely that EEG phases are completely random or PD affects only one of the frequency bands exclusively. Therefore, in this section, we created more realistic simulation data to investigate whether it is the phase or spectral power information that contributes largely to the prediction accuracy of the CRNN model. We created a simulated PD EEG dataset (PDswphase) by swapping the phases of the original PD EEG data with those of one of the HC participants but keeping the spectral power intact (Fig. 8A). This process was repeated 21 times as we copied the phases of one HC participant each time and there are 21 HC participants in total. Similarly, we created another PD EEG dataset (PDswpower) by swapping the power spectrum of PD EEG with the one from a HC participant. In this case, the phase information of the PD EEG was kept the same. This process was also repeated 21 times by utilizing the power spectrum of one HC participant at a time.

**Fig. 8.**
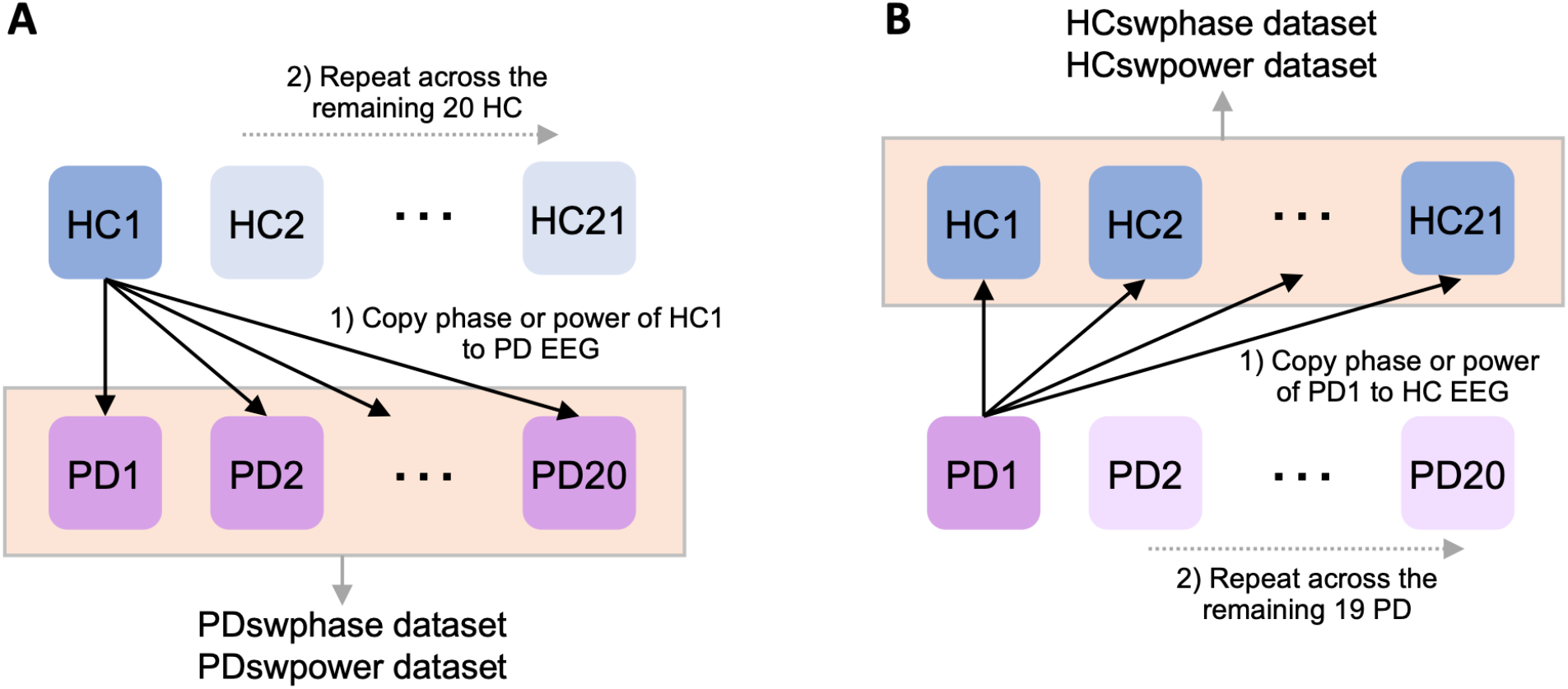
Illustration of the process to create simulated EEG datasets. **(A)** Phase information of HC1’s EEG is copied to the PD EEG samples to create PDswphase dataset. This dataset is used as input to the CRNN model. PDswpower dataset is created by copying spectral power information from HC1 to the PD EEG samples. This process is repeated for the remaining 20 HC participants, creating new PDswphase and PDswpower datasets each time. **(B)** Phase or power information of PD1’s EEG is copied to the HC EEG samples to create HCswphase and HCswpower datasets, respectively. This process is repeated for the remaining 19 PD participants, creating new HCswphase and HCswpower datasets each time.

Fig. 9A shows that phase swapping with the HC participants markedly decreased the model accuracy on the PD EEG samples (9.7 ± 4.1%), indicating that most of the samples were predicted as HC by the phase swapping. The result was consistent, regardless which HC participant’s phase information was used. In contrast, the model accuracy on the PDswpower dataset was in the range of 36.2–62.7% (49.9 ± 8.3%), showing that approximately half of the samples were predicted as HC. Taken together, the results demonstrate that the phase information contributes to the model’s classification much more than the spectral information.

**Fig. 9.**
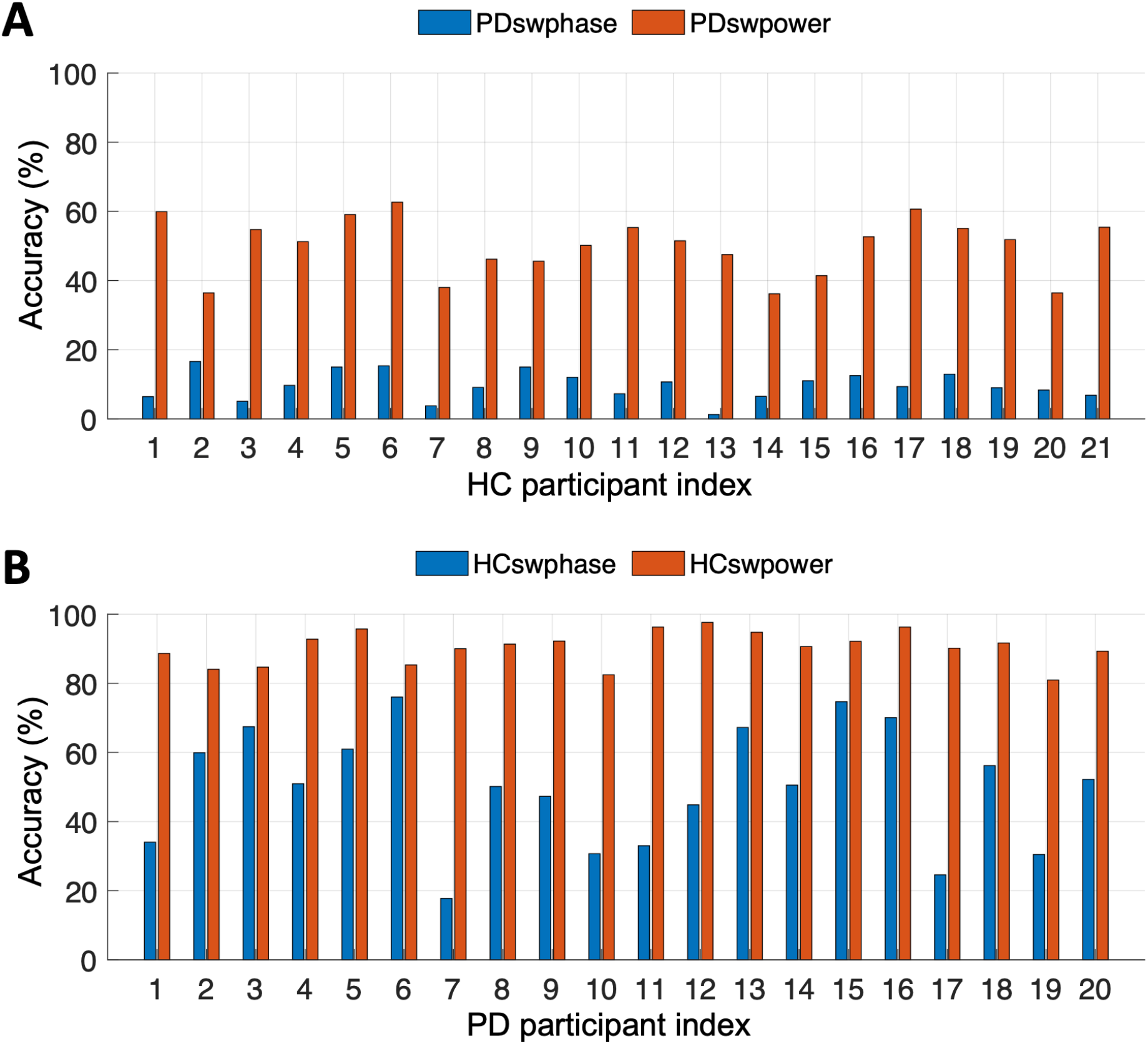
The CRNN model accuracy on the phase- or power-swapped EEG datasets. The x-axis represents the participant index from whom the phases or spectral powers were copied. **(A)** Phases or spectral power were copied from the HC participants to the PD EEG samples. **(B)** Phases or spectral power were copied from the PD participants to the HC EEG samples.

We examined the opposite case where the phases or spectral power of the PD EEG samples were copied to the HC EEG samples. Fig. 9B shows that most of the HC EEG samples (90.4 ± 4.8%) were still correctly predicted when the spectral power was swapped with PD participants. However, when the phase information changed, approximately 50% of the HC samples (50.0 ± 17.1%) were predicted as PD, reconfirming that the phase information contributes more to the high prediction accuracy of the CRNN model compared to the spectral power information.

## 4. Discussion and conclusion

Prior studies have reported that resting-state EEG is altered in PD individuals based on the evidence observed in several time domain, frequency domain, and time-frequency domain features. The main consensus of the findings is a general slowing of the EEG compared to healthy controls (Bočková and Rektor, 2019; Geraedts et al., 2018), which was also reported to be associated with cognitive impairment in PD (Bosboom et al., 2006). Other EEG studies have found that PD is associated with abnormal changes in functional connectivity between cortical regions (Carmona et al., 2017; Silberstein et al., 2005) and nonlinear interaction between lower and higher frequency oscillations (Gong et al., 2020; Swann et al., 2015). These findings highlight that PD is associated with a broad range of linear and nonlinear changes in EEG and the neural basis of the electrophysiological signals underlying the disease is complex and integrative in nature. Therefore, deep learning holds great promise in that it learns good feature representations from complex signals (Craik et al., 2019; Roy et al., 2019).

Applying deep learning, we demonstrated that our model achieves state-of-the-art classification accuracy in the task of discriminating between PD and HC using resting-state EEG. Our model is an end-to-end learning approach where it takes the raw data and performs classification, and therefore it does not require a time-consuming feature extraction step that generally relies on domain knowledge and human expertise and often generates a feature set that insufficiently captures the relevant information in the data. The proposed model is also fundamentally different from the prior EEG studies on functional connectivity in that it looks at relationships across all channels simultaneously through the 1D convolution layers instead of the pairwise relationships. A further advantage of our model is that it is a combination of CNN and RNN, and therefore can capture both temporal and spatial features of EEG that are relevant to the classification task. Our model is a relatively shallow network with two convolutional layers and a GRU layer, yet it outperforms the model consisting of deeper CNNs by a large margin, demonstrating the importance of utilizing sequential features embedded in the raw EEG signals.

Interrogating the proposed model in the simulation studies, we found that the model learns randomized EEG phases as features related to HC. This result is aligned with a recent study (Yi et al., 2017) that investigated the complexity of resting-state cortical activities in PD. It was demonstrated that the EEG of the PD participants had lower permutation entropy (PE) and higher-order index (OI) compared to the HC EEG, suggesting that the resting EEG signals in PD are less complex and more predictable. Similarly, a magnetoencephalography (MEG) study by (Gómez et al., 2011) reported the complexity of the MEG signals is lower in individuals with PD than HC.

We also found that the phase information plays a more significant role than spectral power in our model to discriminate between PD and HC. Phase information has been largely investigated in EEG studies from the perspective of frequency-specific functional connectivity between two brain regions. For example, magnitude squared coherence or wavelet coherence between two EEG channels represents the strength of interaction between the two regions at a frequency of interest. The phase analysis has been much less appreciated in PD studies compared to spectral power, and our findings warrant further investigations of multi-channel phase synchrony in PD.

There are several limitations in our work. Firstly, the small sample size is one of the major deficiencies in this study. Publicly available PD EEG data are scarce, let alone a dataset with two EEG sessions recorded during off- and on-medication states. Also, it is difficult to find public EEG data of healthy individuals who are age-matched to the PD group. Development of a collaborative platform to share data is highly desirable in the future to validate the generalization ability of the proposed model with a larger sample size. Secondly, the interpretability of the proposed model is limited despite we conducted simulation studies to interrogate the model. In recent years, more efforts have been devoted to improving the interpretability of deep learning models and several more advanced models such as an attention-based LSTM (Zhang et al., 2019) have been proposed to improve both the prediction process and interpretability of the time series prediction. This area is being actively researched around the world, and it is expected that such advanced models can provide new valuable insights into abnormal EEG characteristics relevant to PD.

## Data Availability

The data that support the findings of this study are available from the corresponding author, SL, upon reasonable request.

## Authors’ contribution

SL: Conceptualization, Data curation, Formal analysis, Investigation, Methodology, Visualization, Writing - original draft, Writing - review & editing.

RH: Conceptualization, Formal analysis, Methodology, Visualization, Writing - review & editing.

RW: Writing - review & editing.

JZW: Writing - review & editing.

MJM: Conceptualization, Funding acquisition, Investigation, Project administration, Resources, Supervision, Writing - review & editing.

## Acknowledgements

This work is supported by Rina M. Bidin Foundation Fellowship in Research of Brain Treatment (SL), NSERC Canada (MJM), and John Nichol Chair in Parkinson’s Research (MJM).

## Reference

Awan, S.E., Bennamoun, M., Sohel, F., Sanfilippo, F.M., Chow, B.J., Dwivedi, G., 2019. Feature selection and transformation by machine learning reduce variable numbers and improve prediction for heart failure readmission or death. PLoS One 14, e0218760.

Bergstra, J., Bengio, Y., 2012. Random Search for Hyper-Parameter Optimization Yoshua Bengio. J. Mach. Learn. Res. 13, 281–305.

Bind, S., Tiwari, A.K., Sahani, A.K., 2015. A Survey of Machine Learning Based Approaches for Parkinson Disease Prediction. Int. J. Comput. Sci. Inf. Technol. 6, 1648–1655.

Bočková, M., Rektor, I., 2019. Impairment of brain functions in Parkinson’s disease reflected by alterations in neural connectivity in EEG studies: A viewpoint. Clin. Neurophysiol. 130, 239–247.

Bosboom, J.L.W.L.W., Stoffers, D., Stam, C.J.J., van Dijk, B.W.W., Verbunt, J., Berendse, H.W.W., Wolters, E.C.C., 2006. Resting state oscillatory brain dynamics in Parkinson’s disease: an MEG study. Clin. Neurophysiol. 117, 2521–31.

Breiman, L., 2001. Random forests. Mach. Learn. 45, 5–32.

Brown, P., Oliviero, A., Mazzone, P., Insola, A., Tonali, P., Di Lazzaro, V., 2001. Dopamine dependency of oscillations between subthalamic nucleus and pallidum in Parkinson’s disease. J. Neurosci. 21, 1033–8.

Cao, C., Liu, F., Tan, H., Song, D., Shu, W., Li, W., Zhou, Y., Bo, X., Xie, Z., 2018. Deep Learning and Its Applications in Biomedicine. Genomics, Proteomics Bioinforma. 16, 17–32.

Carmona, J., Suarez, J., Ochoa, J., 2017. Brain Functional Connectivity in Parkinson’s disease – EEG resting analysis, in: VII Latin American Congress on Biomedical Engineering CLAIB 2016. Springer, Singapore, pp. 185–188.

Cassani, R., Estarellas, M., San-Martin, R., Fraga, F.J., Falk, T.H., 2018. Systematic review on resting-state EEG for Alzheimer’s disease diagnosis and progression assessment. Dis. Markers 2018.

Christ, M., Braun, N., Neuffer, J., Kempa-Liehr, A.W., 2018. Time Series FeatuRe Extraction on basis of Scalable Hypothesis tests (tsfresh – A Python package). Neurocomputing 307, 72–77.

Craik, A., He, Y., Contreras-Vidal, J.L., 2019. Deep learning for electroencephalogram (EEG) classification tasks: A review. J. Neural Eng. 16, 031001.

Feigin, V.L., Nichols, E., Alam, T., Bannick, M.S., Beghi, E., Blake, N., Culpepper, W.J., Vos, T., et al., 2019. Global, regional, and national burden of neurological disorders, 1990–2016: a systematic analysis for the Global Burden of Disease Study 2016. Lancet Neurol. 18, 459–480.

Geraedts, V.J., Boon, L.I., Marinus, J., Gouw, A.A., Van Hilten, J.J., Stam, C.J., Tannemaat, M.R., Contarino, M.F., 2018. Clinical correlates of quantitative EEG in Parkinson disease: A systematic review. Neurology 91, 871–883.

Gómez, C., Olde Dubbelink, K.T.E., Stam, C.J., Abásolo, D., Berendse, H.W., Hornero, R., 2011. Complexity analysis of resting-state MEG activity in early-stage Parkinson’s disease patients. Ann. Biomed. Eng. 39, 2935–2944.

Gong, R., Wegscheider, M., Mühlberg, C., Gast, R., Fricke, C., Rumpf, J.-J., Nikulin, V.V, Knösche, T.R., Classen, J., 2020. Spatiotemporal features of β-γ phase-amplitude coupling in Parkinson’s disease derived from scalp EEG. Brain awaa400, 1–17.

Groppe, D.M., Bickel, S., Keller, C.J., Jain, S.K., Hwang, S.T., Harden, C., Mehta, A.D., 2013. Dominant frequencies of resting human brain activity as measured by the electrocorticogram. Neuroimage 79, 223–33.

Han, C.X., Wang, J., Yi, G.S., Che, Y.Q., 2013. Investigation of EEG abnormalities in the early stage of Parkinson’s disease. Cogn. Neurodyn. 7, 351–359.

Heijmans, M., Habets, J.G.V., Herff, C., Aarts, J., Stevens, A., Kuijf, M.L., Kubben, P.L., 2019. Monitoring Parkinson’s disease symptoms during daily life: a feasibility study. npj Park. Dis. 5.

Kingma, D.P., Ba, J.L., 2015. Adam: A method for stochastic optimization, in: 3rd International Conference on Learning Representations, ICLR 2015 - Conference Track Proceedings. International Conference on Learning Representations, ICLR.

Koch, M., Geraedts, V., Wang, H., Tannemaat, M., Back, T., 2019. Automated Machine Learning for EEG-Based Classification of Parkinson’s Disease Patients, in: Proceedings - 2019 IEEE International Conference on Big Data, Big Data 2019. Institute of Electrical and Electronics Engineers Inc., pp. 4845–4852.

Lee, S., Hussein, R., McKeown, M.J., 2019a. A deep convolutional-recurrent neural network architecture for Parkinson’s disease EEG classification, in: GlobalSIP 2019 7th IEEE Global Conference on Signal and Information Processing, Proceedings. Institute of Electrical and Electronics Engineers Inc., Ottawa.

Lee, S., Liu, A., Wang, Z.J., McKeown, M.J., 2019b. Abnormal Phase Coupling in Parkinson’s Disease and Normalization Effects of Subthreshold Vestibular Stimulation. Front. Hum. Neurosci. 13, 118.

Mahmoodian, N., Boese, A., Friebe, M., Haddadnia, J., 2019. Epileptic seizure detection using cross-bispectrum of electroencephalogram signal. Seizure 66, 4–11.

Maitín, A.M., García-Tejedor, A.J., Muñoz, J.P.R., 2020. Machine Learning Approaches for Detecting Parkinson’s Disease from EEG Analysis: A Systematic Review. Appl. Sci. 10, 8662.

Makeig, S., Jung, T.-P., Bell, A.J., Sejnowski, T.J., 1996. Independent Component Analysis of Electroencephalographic Data. Adv. Neural Inf. Process. Syst. 8, 145– 151.

Morita, A., Kamei, S., Mizutani, T., 2011. Relationship between slowing of the EEG and cognitive impairment in Parkinson disease. J. Clin. Neurophysiol. 28, 384–387.

Neufeld, M.Y., Blumen, S., Aitkin, I., Parmet, Y., Korczyn, A.D., 1994. EEG frequency analysis in demented and nondemented parkinsonian patients. Dementia 5, 23–8.

Noble, W.S., 2006. What is a support vector machine? Nat. Biotechnol. 24, 1565–1567.

Oh, S.L., Hagiwara, Y., Raghavendra, U., Yuvaraj, R., Arunkumar, N., Murugappan, M., Acharya, U.R., 2018. A deep learning approach for Parkinson’s disease diagnosis from EEG signals. Neural Comput. Appl. 32, 10927–10933.

Oliveira, A.P.S. de, Santana, M.A. de, Andrade, M.K.S., Gomes, J.C., Rodrigues, M.C.A., Santos, W.P. dos, 2020. Early diagnosis of Parkinson’s disease using EEG, machine learning and partial directed coherence. Res. Biomed. Eng. 36, 311–331.

Pereira, C.R., Pereira, D.R., Weber, S.A.T., Hook, C., de Albuquerque, V.H.C., Papa, J.P., 2019. A survey on computer-assisted Parkinson’s Disease diagnosis. Artif. Intell. Med. 95, 48–63.

Poewe, W., Seppi, K., Tanner, C.M., Halliday, G.M., Brundin, P., Volkmann, J., Schrag, A.-E., Lang, A.E., 2017. Parkinson disease. Nat. Rev. Dis. Prim. 3, 17013.

Priori, A., Foffani, G., Rossi, L., Marceglia, S., 2013. Adaptive deep brain stimulation (aDBS) controlled by local field potential oscillations. Exp. Neurol. 245, 77–86.

Roy, Y., Banville, H., Albuquerque, I., Gramfort, A., Falk, T.H., Faubert, J., 2019. Deep learning-based electroencephalography analysis: A systematic review. J. Neural Eng. 16, 501001.

Scandalis, T.A., Bosak, A., Berliner, J.C., Helman, L.L., Wells, M.R., 2001. Resistance training and gait function in patients with Parkinson’s disease. Am. J. Phys. Med. Rehabil. 80, 38–43.

Shah, S.A.A., Zhang, L., Bais, A., 2020. Dynamical system based compact deep hybrid network for classification of Parkinson disease related EEG signals. Neural Networks 130, 75–84.

Shi, X., Wang, T., Wang, L., Liu, H., Yan, N., 2019. Hybrid convolutional recurrent neural networks outperform CNN and RNN in Task-state EEG detection for parkinson’s disease, in: 2019 Asia-Pacific Signal and Information Processing Association Annual Summit and Conference, APSIPA ASC 2019. Institute of Electrical and Electronics Engineers Inc., pp. 939–944.

Silberstein, P., Pogosyan, A., Kühn, A.A., Hotton, G., Tisch, S., Kupsch, A., Dowsey-Limousin, P., Hariz, M.I., Brown, P., 2005. Cortico-cortical coupling in Parkinson’s disease and its modulation by therapy. Brain 128, 1277–91.

Soikkeli, R., Partanen, J., Soininen, H., Pääkkönen, A., Riekkinen, P., 1991. Slowing of EEG in Parkinson’s disease. Electroencephalogr. Clin. Neurophysiol. 79, 159–65.

Su, C., Xu, Z., Pathak, J., Wang, F., 2020. Deep learning in mental health outcome research: a scoping review. Transl. Psychiatry 10, 116(2020).

Swann, N.C., De Hemptinne, C., Aron, A.R., Ostrem, J.L., Knight, R.T., Starr, P.A., 2015. Elevated synchrony in Parkinson disease detected with electroencephalography. Ann. Neurol. 78, 742–750.

Thomas, M., Lenka, A., Kumar Pal, P., 2017. Handwriting Analysis in Parkinson’s Disease: Current Status and Future Directions. Mov. Disord. Clin. Pract. 4, 806–818.

Wainer, J., Cawley, G., 2018. Nested cross-validation when selecting classifiers is overzealous for most practical applications. arXiv.

Winkler, I., Haufe, S., Tangermann, M., 2011. Automatic Classification of Artifactual ICA-Components for Artifact Removal in EEG Signals. Behav. Brain Funct. 7, 30 (2011).

Woo, C.W., Chang, L.J., Lindquist, M.A., Wager, T.D., 2017. Building better biomarkers: Brain models in translational neuroimaging. Nat. Neurosci. 20, 365–377.

Xu, S., Wang, Z., Sun, J., Zhang, Z., Wu, Z., Yang, T., Xue, G., Cheng, C., 2020. Using a deep recurrent neural network with EEG signal to detect Parkinson’s disease. Ann. Transl. Med. 8, 874–874.

Yi, G.-S., Wang, J., Deng, B., Wei, X.-L., 2017. Complexity of resting-state EEG activity in the patients with early-stage Parkinson’s disease. Cogn. Neurodyn. 11, 147–160.

Yuvaraj, R., Rajendra Acharya, U., Hagiwara, Y., 2018. A novel Parkinson’s Disease Diagnosis Index using higher-order spectra features in EEG signals. Neural Comput. Appl. 30, 1225–1235.

Zhang, X., Liang, X., Zhiyuli, A., Zhang, S., Xu, R., Wu, B., 2019. AT-LSTM: An Attention-based LSTM Model for Financial Time Series Prediction, in: IOP Conference Series: Materials Science and Engineering. Institute of Physics Publishing, XuanZhang2019, p. 052037.

