## Supplementary material for "A convolutional-recurrent neural network approach to resting-state EEG classification in Parkinson’s disease"

### Appendix. Supplementary data

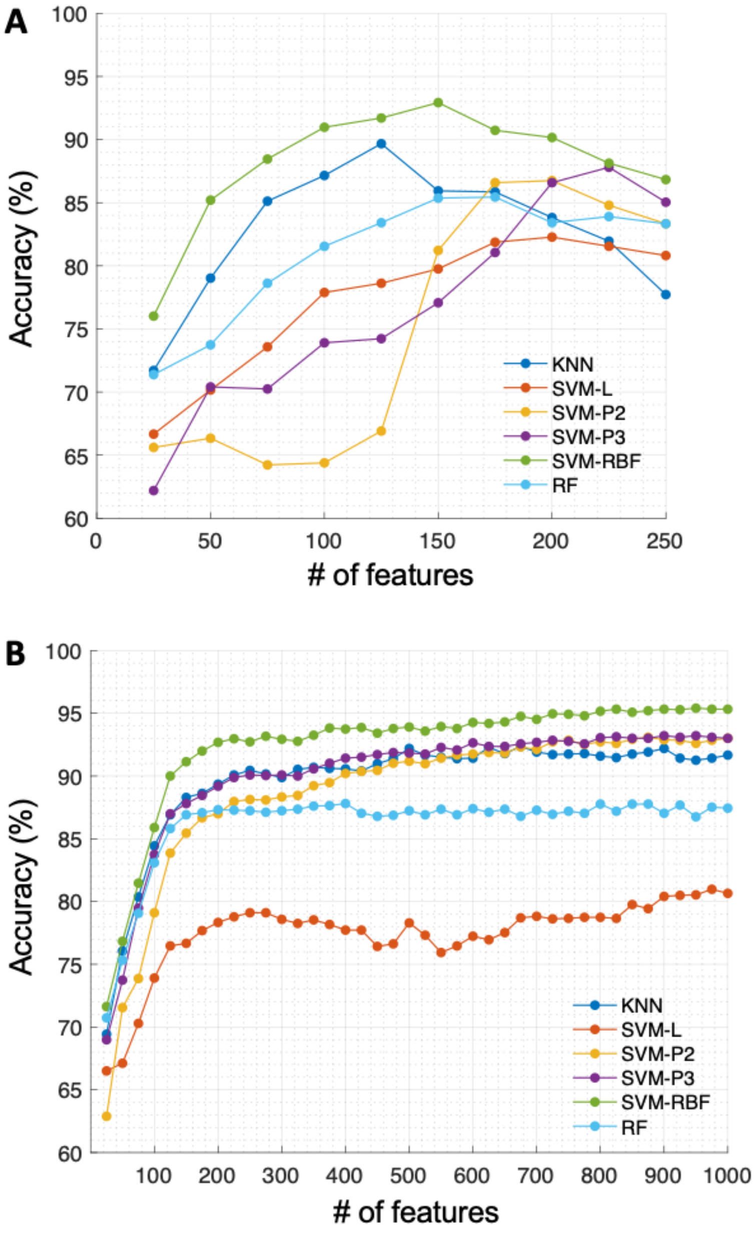


**Fig. S1.** The accuracy of the six different classifiers (KNN, SVM-L, SVM-P2, SVM-P3, SVM-RBF, and RF) as a function of the number of the selected significant features. **(A)** HOS dataset. **(B)** TS dataset.

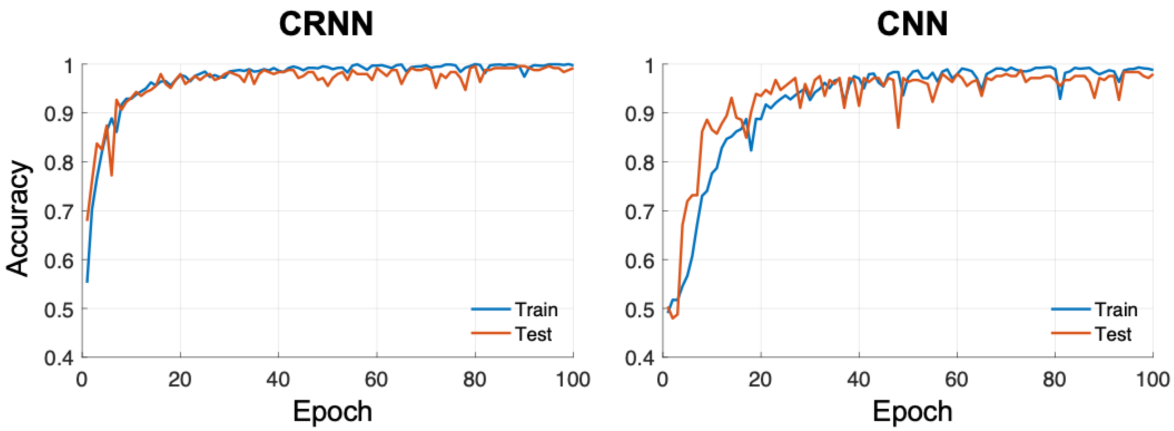


**Fig. S2.** Performance curves of the CRNN (left) and CNN (right) models during the training phase.
